# PREVALENCE AND RISK FACTORS ASSOCIATED WITH MALARIA INFECTION IN CHILDREN ATTENDING LUANGWA DISTRICT HOSPITAL, ZAMBIA; A FACILITY-BASED CROSS-SECTIONAL STUDY

**DOI:** 10.1101/2024.06.23.24309356

**Authors:** Betina Mtonga, Mukumbuta Nawa

**Affiliations:** Department of Biostatistics and Epidemiology, School of Public Health and Environmental Studies, Levy Mwanawasa Medical University, P. O. Box 33991, Lusaka, Zambia

**Keywords:** Malaria infection, children, prevalence, risk factors

## Abstract

**Introduction:** Malaria persists as a major public health issue globally, with sub-Saharan Africa, including Zambia, bearing a disproportionate burden of the disease, necessitating ongoing efforts to combat transmission and improve healthcare access and prevention strategies. This research was aimed at investigating the prevalence and associated risk factors of malaria infection in children attending Luangwa District Hospital, Zambia.

**Methods:** An analytical cross-sectional study was used and the study population included children attending the Out-Patient Department at Luangwa District Hospital. A structured questionnaire and a checklist were used to collect data on malaria infection status, demographic information, and potential risk factors were collected through interviews, medical records review, and laboratory testing. Prevalence rates were calculated using frequencies, and hypothesis tests using the Chi-square test while logistic regression was used to analyse associated factors to malaria infection.

**Results:** A total of 400 children with a mean age of five years participated in the study. Malaria was prevalent in two-thirds of the children (66.2%) with over 57.7% of the infected children presenting with a severe form of malaria and 42.3% having mild disease. 60% had a high malaria parasite density, whereas 40% showed a low density of the parasite. The odds of having malaria were higher for children who did not sleep under Insecticide Treated Nets (ITNs) compared to children who used ITNs (aOR = 24.6, CI = 10.8, 55.7, p-value < 0.001). Furthermore, children whose parents had a secondary-level education compared to parents with no formal education had 3.3 times increased odds of having malaria (aOR = 3.3, CI = 1.09, 9.98, p = 0.034). On the other hand, gender, age of the child, and age of the parent were not significantly associated with malaria infection.

**Conclusion:** This study found a high prevalence of malaria of 66% among children who attended Luangwa District Hospital indicating that Luangwa is still a hotspot with the low transmission province of Lusaka. Factors associated with malaria included not sleeping under ITNs and secondary education. Factors not associated with malaria included gender, age of the child and age of the parent.

## INTRODUCTION

### Background

The global burden of malaria has been declining over the past two decades, however, there has been a sharp increase in the last four years (1). Malaria increased from 229 million in 2019 to 249 million cases in 2022, over 95% of malaria cases and deaths occurred in sub-Saharan Africa (WHO, 2023). Globally, malaria causes 10% of all deaths in children, widespread in tropical and subtropical regions, particularly Sub-Saharan Africa (2). In Zambia, malaria increased from 8.1 million in 2022 to 11.1 million cases in 2023, and the country accounted for 1.4% of the global burden of the disease (3).

In order to accelerate progress towards elimination by 2030, the WHO recommends affected countries and the global malaria community to use the Global Technical Strategy for Malaria Elimination 2016 – 2030 which provides a comprehensive framework to guide countries in their efforts to accelerate progress towards malaria elimination (4). The strategy sets the target of reducing global malaria incidence and mortality rates by at least 90% by 2030 (4). Zambia has implemented and continues to implement the fight against malaria using the National Malaria Elimination Strategic Plan 2017 – 2025 which has locally adapted the GTS strategies of malaria control in high-burden areas and elimination strategies in low-transmission areas (5, 6). The burden of malaria in Zambia is highest in northern parts of the country such as Northern, Luapula and Muchinga provinces, which receive the highest rainfall and the malaria incidence is over 500 cases per 1000 population (7). These are followed by the provinces in the middle of the country such as Eastern, Copperbelt, Central North-Western and Western provinces where malaria incidence ranges from 50 to 499 cases per 1000 population (8, 9). The southern parts of the country including Southern and Lusaka provinces have the lowest burden of malaria in Zambia with incidence cases below 50 per 1000 population partly due to lower rainfall and droughts experienced in these areas (7). Implementation of malaria interventions such as mass distribution of Insecticide-Treated Nets (ITNs) and Indoor Residual Spraying is done throughout the country whilst elimination interventions are only done in selected areas with low transmission (6).

In low transmission areas of Lusaka province, Luangwa district where the Luangwa River makes a confluence with the Zambezi River is a low-lying area with a hot climate and swampy landscapes which support mosquito habitats and is a malaria hotspot in the province (10, 11). This study aimed to assess the prevalence and associated risk factors among children attending Luangwa District Hospital which is a first-level referral hospital in the district.

## METHODOLOGY

### Study design

A facility-based analytical cross-sectional study design was conducted from February to March 2024, at Luangwa District Hospital involving determination of malaria prevalence and its associated risk factors among children attending the health facility.

### Study Site

The study was conducted at Luangwa District Hospital, situated in Luangwa District, Zambia, within the Lusaka province. According to the 2020 Zambian Census, the district’s population was 31,007 people. Luangwa District is bordered by two rivers: The Luangwa River to the east, forming the boundary with Eastern Province and the Zambezi River to the south, serving as the border with Zimbabwe. The district’s residents primarily engage in farming and fishing as their main occupations, facilitated by the surrounding water bodies, which also create conducive environments for mosquito breeding.

### Study Population

The study included children attending medical services from the Out-Patient Department at Luangwa District Hospital from February to March 2024.

### Eligibility criteria

#### Eligibility Criteria

1. Age: Children under the age of 12 years.
2. Geographic Location: Children residing in Luangwa district, Zambia.
3. Health Status: Children without underlying health issues.
4. Parental Consent: Children whose parents or legal guardians have provided informed consent for their participation in the study.

#### Exclusion Criteria

Children who had other known underlying conditions and those who were too sick were excluded

### Sample Size Determination

Preliminary findings showed that an average of 483 patients visit the Out-Patient Department at Luangwa District Hospital per week and the sample size was thus based on this figure. Therefore, the formula below was used to determine the sample size since the population size was known. The study was done at 95% Cl and powered at 80%.

Where:

- *n* = the required sample size
- Z = the Z-score corresponding to the desired confidence level (e.g., 1.96 for a 95% confidence level)
- *p* = the estimated proportion of the population with the characteristic of interest (50%)
- e = the margin of error (expressed as a proportion)

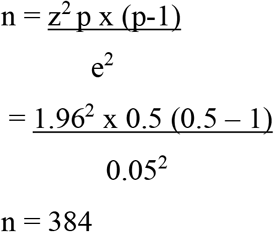

Therefore, the minimum sample size for this study was 400 after inflating the sample with a 10% non-response rate.

### Data analysis

The collected was collected using a questionnaire using random sampling among attendees using a questionnaire administered by trained research assistants using the local language (Nyanja) and was entered into Microsoft Excel. Data cleaning and statistical analysis were done in STATA version 16. Frequencies were done to determine the prevalence of malaria, and Chi-square analysis and logistic regression analysis were performed to identify factors associated with malaria infection at a confidence level of 95%. Light microscopy was used to measure malaria parasitemia.

### Ethical Considerations

The study was approved by the Lusaka Apex University Research Ethics Committee (IRB 00001131, FWA 0029892 and Approval Reference No. 00741-24). Consent was attained from the parents and guardians of the participating children. Anonymity, as well as confidentiality, were observed at all times using codes on the questionnaires of the participants other than names.

## RESULTS

### Participant’s baseline characteristics

A total of 400 children participated in this study, majority 251 (62.7%) were boys compared to girls 149 (37.3%), about 80 (20%) were aged below one year whilst 200 (50%) were aged one to five years and those above five years were 120 (30%). The baseline characteristics of study participants are summarized in Table 1.

**Table 1:**
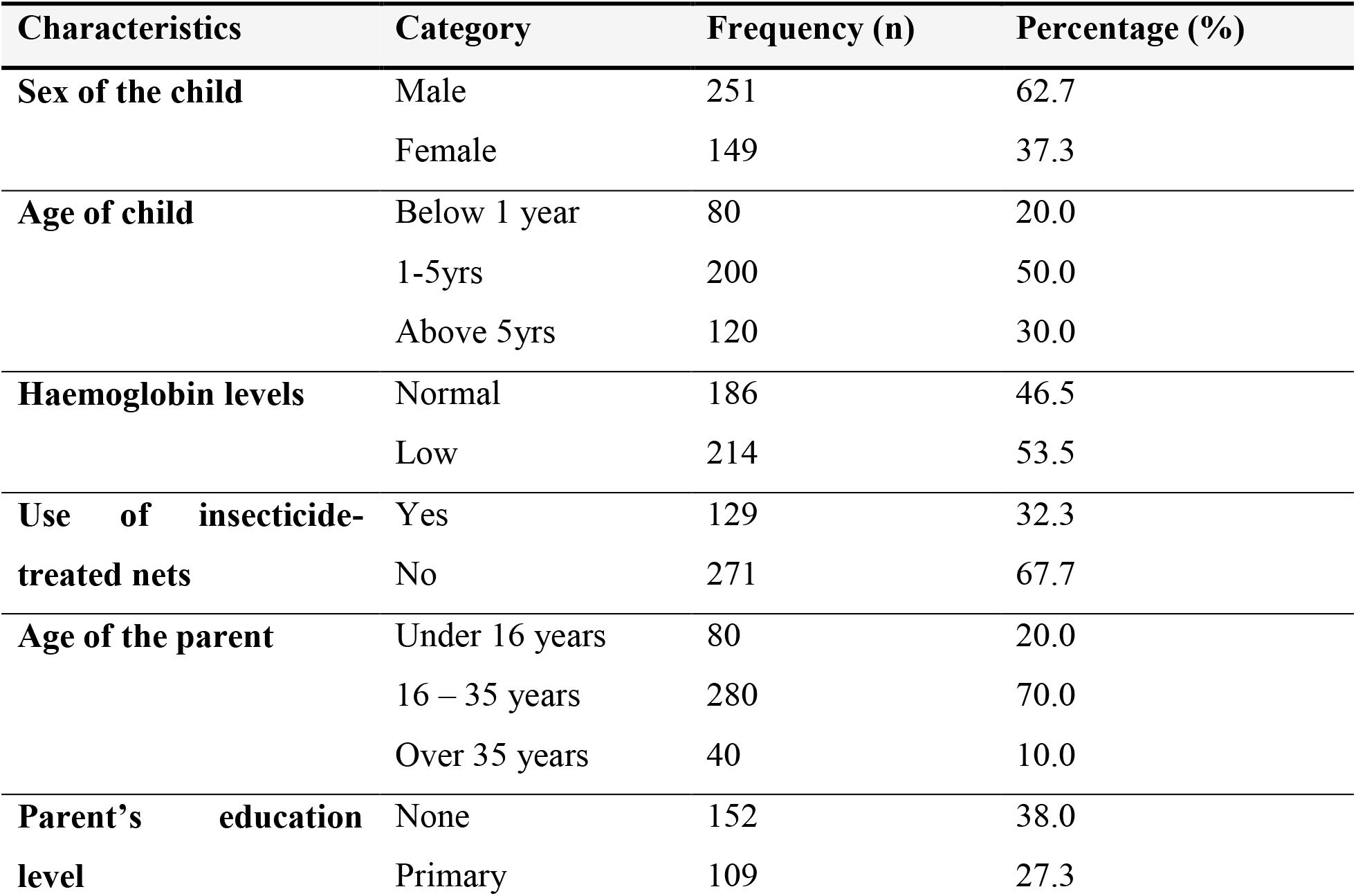
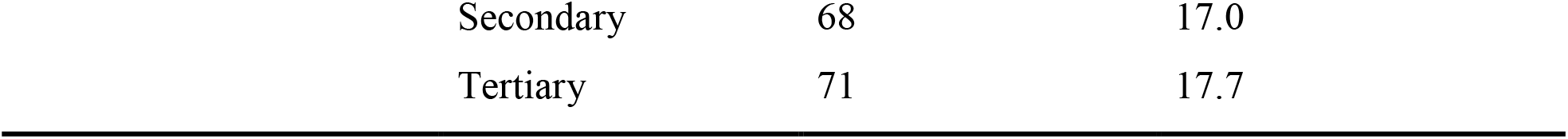
Socio- demographics.

| **Characteristics** | **Category** | **Frequency (n)** | **Percentage (%)** |
| --- | --- | --- | --- |
| **Sex of the child** | Male | 251 | 62.7 |
|  | Female | 149 | 37.3 |
| **Age of child** | Below 1 year | 80 | 20.0 |
|  | 1-5yrs | 200 | 50.0 |
|  | Above 5yrs | 120 | 30.0 |
| **Haemoglobin levels** | Normal | 186 | 46.5 |
|  | Low | 214 | 53.5 |
| **Use of insecticide-treated nets** | Yes | 129 | 32.3 |
|  | No | 271 | 67.7 |
| **Age of the parent** | Under 16 years | 80 | 20.0 |
|  | 16 – 35 years | 280 | 70.0 |
|  | Over 35 years | 40 | 10.0 |
| **Parent's education level** | None | 152 | 38.0 |
|  | Primary | 109 | 27.3 |

|  |  |  |
| --- | --- | --- |
| Secondary | 68 | 17.0 |
| Tertiary | 71 | 17.7 |

### Prevalence of malaria among participants

Malaria was present in two-thirds of the children 66.2% (265), with over half of the infected children presenting with a severe form of malaria 153 (57.7%), and 42.3% (112) having mild disease. Among the infected children, 60% (159) had a high malaria parasite density, whereas 40% (106) showed a low density of the parasite, Table 2 shows the breakdown of malaria, severity of symptoms and parasite density.

**Table 2:**
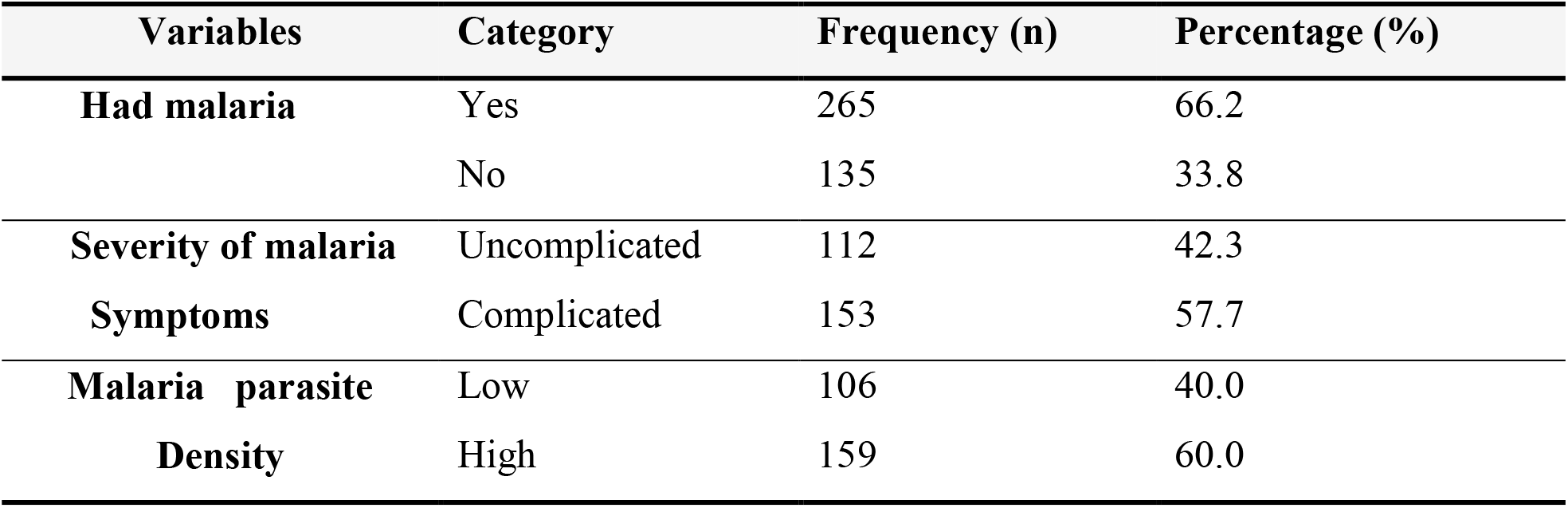
Malaria Prevalence and Severity.

### Factors associated with the prevalence of malaria

Results in Table 3 show that the prevalence of malaria was significantly different between male and female children (55.4% vs. 84.6%, p-value < 0.001), Similarly, the proportion of malaria cases significantly differed between those who used ITN and those who did not (27.1% vs. 84.9%, p-value < 0.001) and across different levels of education (p-value = 0.002).

**Table 3:** Outcome by Independent Variables.

| Variable | Category | Had Malaria infection |  | P value* |
| --- | --- | --- | --- | --- |
|  |  | Yes, N = 265<br>n (%) | No, N = 135<br>n (%) |  |
| **Sex of the child** | Male | 139 (55.4) | 112 (44.6) | **< 0.001** |
|  | Female | 126 (84.6) | 23 (15.4) |  |
| **Age of child** | Below 1 year | 51 (63.7) | 29 (36.3) | 0.855 |
|  | 1-5yrs | 133 (66.5) | 67 (33.5) |  |
|  | Above 5yrs | 81 (67.5) | 39 (32.5) |  |
| **Haemoglobin levels** | Normal | 112 (60.2) | 74 (39.8) | **0.017** |
|  | Low | 153 (71.5) | 61 (28.5) |  |
| **Use of insecticide-treated nets** | Yes | 35 (27.1) | 94 (72.9) | **< 0.001** |
|  | No | 230 (84.9) | 41 (15.1) |  |
| **Age of the parent** | Under 16 years | 51 (63.7) | 29 (36.3) | 0.866 |
|  | 16 – 35 years | 187 (66.8) | 93 (33.2) |  |
|  | Over 35 years | 27 (67.5) | 13 (32.2) |  |
| **Parent's education level** | None | 106 (69.7) | 46 (30.3) | **0.002** |
|  | Primary | 63 (57.8) | 46 (42.2) |  |
|  | Secondary | 56 (82.4) | 12 (17.6) |  |
|  | Tertiary | 40 (56.3) | 31 (43.6) |  |

\*Chi-Square Test of Association

The study found that those who did not use ITNs were at an increased odds of having malaria compared to children who slept under (aOR = 24.6, CI = 10.8, 55.7, p-value < 0.001). Furthermore, children whose parents had a secondary-level education compared to parents with no formal education had increased odds of having malaria (aOR = 3.3, CI = 1.09, 9.98, p = 0.034). On the other hand, gender, age of the child, and age of the parent had no significant effect on the odds of malaria infection at multivariable analysis (all p-values > 0.05). Table 4 summarises the factors associated with malaria infection.

**Table 4:** Factors Associated with Prevalence of Malaria Infection.

| Variable | uOR | 95% CI | P value | aOR | 95% CI | P value |
| --- | --- | --- | --- | --- | --- | --- |
| **Sex of the child** |  |  |  |  |  |  |
| Male | **Ref** |  |  | **Ref** |  |  |
| Female | 4.41 | 2.65, 7.35 | **<0.001** | 0.99 | 0.46, 2.14 | 0.973 |
| **Age of child** |  |  |  |  |  |  |
| Below 1 year | **Ref** |  |  | **Ref** |  |  |
| 1-5yrs | 1.13 | 0.66, 1.94 | 0.662 | 3.05 | 0.52, 17.7 | 0.215 |
| Above 5yrs | 1.18 | 0.65, 2.14 | 0.584 | 2.65 | 0.66, 10.7 | 0.172 |
| **Use of insecticide-treated nets** |  |  |  |  |  |  |
| Yes | **Ref** |  |  | **Ref** |  |  |
| No | 15.01 | 9.04, 25.1 | **<0.001** | 24.6 | 10.8, 55.7 | **<0.001** |
| **Age of the parent** |  |  |  |  |  |  |
| Under 16 years | **Ref** |  |  | **Ref** |  |  |
| 16 – 35 years | 1.14 | 0.68, 1.92 | 0.613 | 1.07 | 0.29, 3.87 | 0.923 |
| Over 35 years | 1.18 | 0.53, 2.64 | 0.685 | — | — | — |
| **Parent's education level** |  |  |  |  |  |  |
| None | **Ref** |  |  | **Ref** |  |  |
| Primary | 0.59 | 0.36, 0.99 | **0.047** | 0.95 | 0.40, 2.33 | 0.927 |
| Secondary | 2.02 | 0.99, 4.13 | 0.052 | 3.30 | 1.09, 9.98 | **0.034** |
| Tertiary | 0.56 | 0.31, 1.00 | 0.051 | 1.27 | 0.53, 3.03 | 0.593 |

**uOR** = unadjusted odds ratio, **aOR** = adjusted odds ratio, **CI** = Confidence interval

## DISCUSSION

This study set out to find the prevalence and risk factors associated with malaria infection among children aged 0 – 12 years attending a first-level hospital in Luangwa district which is a malaria hotspot within Lusaka province with low malaria transmission. The findings indicate that about two-thirds of the children who attended this hospital had malaria infection. This high level of malaria infections among children is concerning and a reflection of community infections taking place in the district. This was not a population survey to reflect the community-level prevalence, which is expected to be lower because our study population comprised of children who were sick and seeking treatment at the hospital whilst, in the community, not all the children were sick. A recent country-wide survey found that prevalence among under-five children in Zambia was 29% which is lower than our study findings (12). Specifically, in Lusaka province, the Malaria Indicator Survey 2021 found a prevalence of 3.3% which was way below the 66.2% that this study found; moreover, this was for the whole province which is a low transmission whilst our study was for one district which is a malaria hotspot (12). Further, the MIS only tests malaria in under-five children whilst our study tested even older children. Previous studies have shown that children aged 1 year to five years tend to have higher malaria infections compared to children below one year (11, 12). Our study which included children up to 12 years is therefore expected to have a lower estimate since older children are known to develop partial immunity to malaria, but the main reason for the higher prevalence is the selection bias of sick children seeking treatment (13). There were no recent studies specifically in Luangwa district, however, an earlier study had found a prevalence of 9.7% in under-five children but this study was done about 14 years before our study and it was a community-based survey and therefore cannot be compared with our study (14). Nonetheless, our study affirms that Luangwa district is still a malaria hotspot within Lusaka province which is a low transmission area in Zambia.

The predominance of males (63%) in the study population compared to female children may suggest potential gender-related health disparities or differences in healthcare-seeking behaviour, however, another study in Zambia specifically on health-seeking behaviour did not find any significant differences between boys and girls for common childhood illnesses (15). The sex of the child was not statistically associated with the risk of malaria, this is similar to other studies on malaria in other countries such as Nigeria, a country with the highest malaria burden in sub-Saharan Africa (16). In this study, older children were found to have higher odds of malaria compared to children below one year but this was not statistically significant, other studies found that children older than one year were more at risk of malaria compared to those below one year (16). A recently published scoping review of studies in sub-Saharan Africa on risk factors for malaria has shown that the association between age and malaria was inconsistent (17). It is however, established that older children above five years and adults are less likely to have clinical malaria as they develop partial immunity and as such tend to have subclinical parasitaemia (13). The age of the parent was not associated with increased odds of the child having malaria, but the parent’s education was associated with the risk of malaria; children whose parents had secondary education were associated with higher odds of malaria compared to those with no education, however, this is contrary to other studies which found that higher education was associated with less risk of malaria as they can access preventive interventions (18). Similarly, those with higher economic status tend to have better housing and nutrition for their children and therefore less risk of malaria (18). Our study may have found this seemingly counter-intuitive result that those with secondary education had a higher risk of malaria because of the nature of the study which was facility-based. Better health-seeking behaviour and access to health services are skewed towards those who are more educated compared to those with no education (15). Another study multi-centre study in 10 African countries found that higher education was associated with better health-seeking behaviour than those with less education (19).

Additionally, our study found a significant association between the use of interventions like Insecticide-Treated Nets (ITNs) and reduced odds of malaria infection among children. This is consistent with existing literature on ITNs reducing malaria transmission, a recent systematic review and meta-analysis of 11 studies in Ethiopia found that the use of ITNs was associated with lower odds of malaria (20). ITN use is protective against malaria for individuals using them, especially in Africa where the primary vectors are endophagic and anthropophilic (21). Other factors that may show reduced effects of ITNs include the changes in the feeding behaviour of primary mosquitoes where there is high coverage (22). Other threats to ITN effectiveness include emerging resistance to commonly used insecticides such as pyrethroids by primary mosquito vectors (23). In our study site, ITNs are found to still be effective despite these emerging threats.

## CONCLUSION

This study found a high prevalence of malaria of 66% among children who attended Luangwa District Hospital indicating that Luangwa is still a hotspot with the low transmission province of Lusaka. Factors associated with malaria included not sleeping under ITNs and secondary education. Factors not associated with malaria included gender, age of the child and age of the parent.

## RECOMMENDATIONS

The study recommends the continued use of insecticide-treated bed nets (ITNs) in the fight against malaria in Luangwa district which is a hotspot for malaria transmission in a province of low malaria transmission.

## LIMITATIONS

The study employed a facility-based cross-sectional design which does not reflect the prevalence of malaria at the community level but among those who are seeking treatment at the district hospital, it cannot therefore be generalized to the entire district. Further, the cross-sectional design does not establish causal relationships between risk factors and malaria infection. Longitudinal or prospective studies would be required to explore temporal relationships.

## Data Availability

All data produced in the present study are available upon reasonable request to the authors

## Notes

### Competing Interest Statement

The authors have declared no competing interest.

### Funding Statement

This study did not receive any funding

### Author Declarations

Lusaka Apex Medical University Research Ethics Committee - IRB 00001131, FWA 0029892 and Approval Reference No. 00741-24.

